# Elevated fecal mitochondrial DNA from symptomatic norovirus infections suggests potential health relevance of human mitochondrial DNA in fecal source tracking

**DOI:** 10.1101/2022.02.24.22271477

**Authors:** Kevin J. Zhu, Brittany Suttner, Jackie Knee, Drew Capone, Christine L. Moe, Christine E. Stauber, Kostas T. Konstantinidis, Thomas E. Wallach, Amy J. Pickering, Joe Brown

## Abstract

An end goal of fecal source tracking (FST) is to provide information on risk of transmission of waterborne illnesses associated with fecal contamination. Ideally, concentrations of FST markers in ambient waters would reflect exposure risk. Human mtDNA is an FST marker that is exclusively human in origin and may be elevated in feces of individuals experiencing gastrointestinal inflammation. In this study, we examined whether human mtDNA is elevated in fecal samples from individuals with symptomatic norovirus infections using samples from the US, Mozambique, and Bangladesh. We quantified hCYTB484 (human mtDNA) and HF183/BacR287 (human-associated *Bacteroides*) FST markers using droplet digital PCR. We observed the greatest difference in concentrations of hCYTB484 when comparing samples from individuals with symptomatic norovirus infections versus individuals without norovirus infections or diarrhea symptoms: log_10_ increase of 1.42 in US samples (3,820% increase, *p*-value = 0.062), 0.49 in Mozambique (308% increase, *p*-value = 0.061), and 0.86 in Bangladesh (648% increase, *p*-value = 0.035). We did not observe any trends in concentrations of HF183/BacR287 in the same samples. These results suggest concentrations of fecal mtDNA increase during symptomatic norovirus infection and that mtDNA in environmental samples may represent an unambiguously human source-tracking marker that correlates with enteric pathogen exposure risk.

## Introduction

Fecal source tracking (FST) aims to detect fecal contamination in environmental samples and identify the source using a variety of chemical and biological methods. Method validation studies to date have demonstrated FST markers targeting human-associated microbial DNA to have variable sensitivity (true positive rate) and specificity (true negative rate) across geographies ^1–5^. Human mitochondrial DNA (mtDNA) markers, having been demonstrated to have high sensitivity ^6–9^ and high specificity ^6–9^ across varying geographies, may complement the use of microbial FST targets, especially in settings or environmental matrices where other FST markers have not been previously validated ^9^.

In addition to retaining high sensitivity and specificity, an ideal FST marker would also convey information about the risk associated with detected fecal contamination: increasing concentration of FST markers in environmental samples should indicate increasing risk of gastrointestinal and other waterborne illnesses associated with exposure ^10^. This increasing risk may be due to an increase in fecal input generally increasing chances of pathogens being present or due to a fecal source present with particularly high concentrations of infectious pathogens. However, such a marker has not yet been identified. mtDNA FST markers differ from microbial markers because they target host DNA instead of host-associated microbial DNA. The main cellular sources of fecal mtDNA are thought to be intestinal epithelial cells (IEC) ^6,7^ and white blood cells (leukocytes) ^6,11^. IECs constitute the intestinal epithelium that prevents the entry of harmful substances into the body while selectively allowing entry of beneficial nutrients. Leukocytes transmigrate into the intestinal lumen during enteric infections. Because of these origins, fecal mtDNA concentrations may exhibit baseline levels during homeostasis (e.g., IEC shedding to balance IEC proliferation) with elevated levels during inflammatory events (e.g., infection triggering leukocyte transmigration, increased apoptosis, IEC extrusion and shedding) ^12,13^.

A key assumption often used when assessing risk associated with fecal indicators is that concentrations of fecal indicators covary with concentrations of sewage present and, therefore, concentrations of pathogens. If concentrations of mtDNA FST markers increase during cases of enteric infections, specifically in symptomatic cases where vomiting and/or diarrhea facilitates the shedding of pathogens, mtDNA markers may advance the capabilities of FST markers by providing risk information beyond the assumed covariance between concentrations of indicator and pathogen. Associations between mtDNA FST markers and enteric infections have not yet been studied, and it is unknown whether concentrations of fecal mtDNA are indicative of symptomatic enteric infections.

The aim of this study was to investigate whether concentrations of a human mtDNA FST marker are higher in feces from individuals with symptomatic norovirus infections than feces from individuals without. We used archived fecal samples from participants in studies conducted in the US, Mozambique, and Bangladesh, and compared fecal mtDNA concentrations across three groups: (1) no detected enteric infection and no diarrhea, (2) norovirus infection and no diarrhea, and (3) norovirus infection and diarrhea. We hypothesized that concentrations of fecal mtDNA will be highest in feces from symptomatic norovirus infections versus those from individuals with asymptomatic norovirus infections or no enteric infections.

## Materials and Methods

### Feces Samples

We obtained human fecal samples from three different studies conducted in the US, Mozambique, and Bangladesh. We first investigated the US samples, using pairs of one pre- and one post-challenge sample per subject from a norovirus Genogroup I (GI) challenge study in which norovirus-spiked oysters were used as the intentional exposure ^14^. These pre- and post-challenge pairs were from six subjects who developed asymptomatic norovirus infections and five subjects who developed symptomatic norovirus infections (Table 1). In the challenge study, symptoms (chills, cramping, diarrhea, fatigue, fever, headache, myalgia, nausea, vomiting, white blood cell shift) were recorded during the challenge period and follow-up visits ^14^. To be classified as symptomatic, a subject had to have at least one of the above symptoms, with fever requiring at least one other associated symptom.

**Table 1.** Sample frame for this study.

| Study population | Individuals from which samples were collected | Ages of individuals | enteric<sup>-</sup>asymptomatic | noro<sup>+</sup>asymptomatic | noro<sup>+</sup>symptomatic |
| --- | --- | --- | --- | --- | --- |
| US <sup>a</sup> | 11 | Adult (18 to 50 years of age) | 11 | 6 | 5 |
| Mozambique | 66 | Children (< 4 years of age) | 26 | 29 | 11 |
| Bangladesh | 120 | Children (< 5 years of age) | 49 | 68 | 3 |

<sup>a</sup> All US samples were part of paired samples (pre-challenge and post-challenge) from 11 individuals. 6 individuals did not develop symptoms; 5 individuals did develop symptoms.

Following initial results from the US samples (Figure S1), we expanded the analysis to include archived fecal samples from two other studies: 1) a cross-sectional study of child (under four years of age) enteric infections in urban Maputo, Mozambique ^15,16^ and 2) an experimental trial evaluating the effect of passive chlorination devices at shared water points on child (under five years of age) diarrhea prevalence in urban Bangladesh ^17,18^. We classified the fecal samples using the following criteria: 1) no enteric pathogens detected and from individuals with no reported diarrhea (hereafter referred to as enteric^-^asymptomatic), 2) norovirus GI/GII detected and from individuals with no reported diarrhea (noro^+^asymptomatic), 3) norovirus GI/GII detected, and from individuals with reported diarrhea (noro^+^symptomatic). Detection of enteric pathogens in the archived Mozambique and Bangladesh samples was determined by the Luminex (Austin, Texas, US) xTAG® Gastrointestinal Pathogen Panel RUO (GPP) in previous studies ^15,18^. The GPP detects the nucleic acid markers of 15 bacterial, viral, and parasitic enteric pathogens, including norovirus GI/GII with a limit of detection for norovirus GI/GII on the order of 10^6^ genome copies/gram of feces ^19^. There were low numbers of norovirus-positive feces in the Mozambique and Bangladesh samples, and feces positive for norovirus were often positive for another GPP target ^15,18^. Because of this, we included norovirus-positive feces that were positive for additional pathogen(s) in the noro^+^asymptomatic and noro^+^symptomatic groups. We identified enteric^-^asymptomatic samples by selecting feces that were negative for all Luminex GPP targets. Reported diarrhea in the Bangladesh and Mozambique studies was based on caregiver-reported diarrhea criteria of ≥3 loose or watery feces in a 24-hr period with a 1-week recall period. Because the Bangladesh ^17^ and Mozambique ^15^ studies observed low prevalence of caregiver-reported diarrhea, we were limited in the number of noro^+^symptomatic samples we could examine (Table 1).

### DNA Extraction and ddPCR

Prior to DNA extraction, we stored fecal samples at −80°C. For US samples, we performed DNA extractions using 0.1 g of fecal sample and the MO BIO PowerSoil® kit (Carlsbad, CA, USA) following manufacturer’s instructions. For Mozambique and Bangladesh samples, we performed DNA extractions using 0.1 g of fecal sample and the Qiagen QIAamp® 96 PowerFecal QIAcube® HT Kit automated on the Qiagen QIAcube® HT platform (Hilden, Germany) following manufacturer’s instructions, using soil grinding SK38 bead tubes (Bertin Corp., Rockville, MD, USA) containing 650 µl pre-warmed Buffer PW1 and homogenizing the bead tubes on a vortexer for 10 minutes. Following DNA extraction, we stored all extracts at − 80°C until analysis. We quantified hCYTB484 ^9^ and HF183/BacR287 ^20^ markers through droplet digital PCR (ddPCR) on Bio-Rad QX200™ Droplet Digital™ PCR (Hercules, CA, USA) using methods developed previously ^9^ and normalized marker concentrations to nanograms of double stranded DNA (ng dsDNA) as measured by Qubit 3.0 Fluorometer with Qubit High Sensitivity DNA kits (ThermoFisher Scientific, Waltham, MA, USA) to account for differences in moisture content between feces and any potential differential recovery between kits ^21^. Results of biological replicates for a subset of samples are in supporting information (Table S1). Minimum Information for Publication of Quantitative Digital PCR Experiments ^22^ is included in the supporting information (Table S2). For both assays, we classified samples as not detected if amplification was below our analytical limit of detection of three positive partitions per ddPCR well ^9^. For the analytical lower limit of quantification, we used previously established assay-specific limits ^9^.

### Data Analysis

Because the US samples were collected pre- and post-challenge from each subject, we applied the Wilcoxon signed rank paired test. For the Mozambique and Bangladesh sample sets (cross-sectional data), we used the Kruskal-Wallis test, followed with the Dunn test with Benjamini-Hochberg adjustment. We calculated effect sizes for log_10_ transformed concentrations through a difference in means approach using Cohen’s *d*, the difference between the two means divided by the pooled standard deviation. To compare the relative influences of potential confounders, we fitted a generalized linear model (GLM) using a Gaussian identity function to the Mozambique and Bangladesh sample sets using reported diarrhea and norovirus GI/GII detected/not detected (as determined by the GPP) as the independent variables and log_10_ values of hCYTB484 normalized to ng of dsDNA as the dependent variable while adjusting for number of pathogens detected (as determined by the GPP), sex, age (continuous, number of months), and study population (Mozambique or Bangladesh). More information on model fitting can be found in the supporting information. We performed data analyses in R version 4.0.1.

## Results and Discussion

We detected hCYTB484 above quantifiable levels in 100% of the samples in this study and found increases in hCYTB484 in samples from symptomatic norovirus infections. We observed the largest differences in median hCYTB484 copies / ng dsDNA between the enteric^-^asymptomatic and noro^+^symptomatic groups (Figure 1, Table 2): a log_10_ increase of 1.42 for US samples (3,820% increase, *p*-value of 0.062, effect size = 4.3), 0.49 increase for Mozambique samples (308% increase, *p*-value of 0.061, effect size = 0.70), and 0.86 increase for Bangladesh samples (648% increase, *p*-value of 0.035, statistically significant at α = 0.05 level, effect size = 1.5). The larger effect sizes between enteric^-^asymptomatic and noro^+^symptomatic versus enteric^-^asymptomatic and noro^+^asymptomatic across all three countries (Table 2) suggest that fecal mtDNA concentrations are higher in symptomatic norovirus infections than in asymptomatic norovirus infections (effect sizes calculated as the difference between means normalized to the pooled standard deviation). To investigate what variables influenced fecal mtDNA concentrations, we standardized the GLM regression coefficients to account for different units of measurements and variances of each variable. The standardized GLM regression coefficients (Table S3) show reported diarrhea (0.16, 95% CI: 0 – 0.32, *p*-value = 0.045), norovirus detected (0.12, 95% CI: − 0.16 – 0.39, *p*-value = 0.38), age in months (−0.12, 95% CI: −0.28 – 0.03, *p*-value = 0.12), and study population (Bangladesh or Mozambique) (0.19, 95% CI: −0.36 – −0.03, *p*-value = 0.018) as having the largest magnitudes. However, only reported diarrhea and study population had *p*-values < 0.05 (0.045 and 0.018, respectively). Comparison of the standardized regression coefficients after adjusting for other potential biological confounders suggest that, of the variables tested, diarrhea and study population had the largest influences on fecal mtDNA concentrations.

**Table 2.**
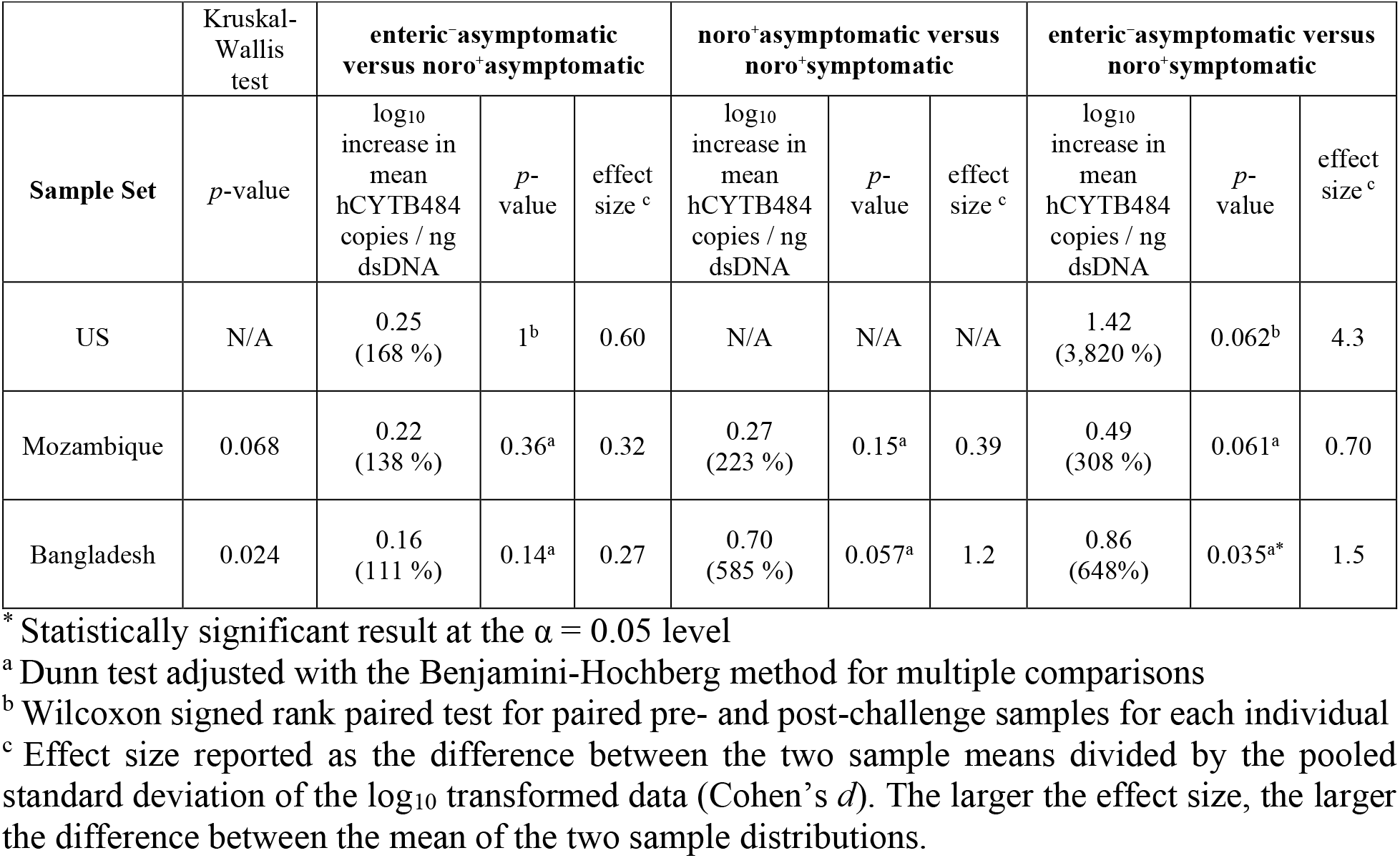
Comparison of log_10_ human mtDNA copies normalized to ng of dsDNA amongst the different health status groups.

| Sample Set | Kruskal-Wallis test | enteric<sup>-</sup>asymptomatic versus noro<sup>+</sup>asymptomatic |  |  | noro<sup>+</sup>asymptomatic versus noro<sup>+</sup>symptomatic |  |  | enteric<sup>-</sup>asymptomatic versus noro<sup>+</sup>symptomatic |  |  |
| --- | --- | --- | --- | --- | --- | --- | --- | --- | --- | --- |
|  | *p*-value | log<sub>10</sub> increase in mean hCYTB484 copies / ng dsDNA | *p*-value | effect size <sup>c</sup> | log<sub>10</sub> increase in mean hCYTB484 copies / ng dsDNA | *p*-value | effect size <sup>c</sup> | log<sub>10</sub> increase in mean hCYTB484 copies / ng dsDNA | *p*-value | effect size <sup>c</sup> |
| US | N/A | 0.25 (168 %) | 1<sup>b</sup> | 0.60 | N/A | N/A | N/A | 1.42 (3,820 %) | 0.062<sup>b</sup> | 4.3 |
| Mozambique | 0.068 | 0.22 (138 %) | 0.36<sup>a</sup> | 0.32 | 0.27 (223 %) | 0.15<sup>a</sup> | 0.39 | 0.49 (308 %) | 0.061<sup>a</sup> | 0.70 |
| Bangladesh | 0.024 | 0.16 (111 %) | 0.14<sup>a</sup> | 0.27 | 0.70 (585 %) | 0.057<sup>a</sup> | 1.2 | 0.86 (648%) | 0.035<sup>a*</sup> | 1.5 |

\* Statistically significant result at the $\alpha = 0.05$ level
<sup>a</sup> Dunn test adjusted with the Benjamini-Hochberg method for multiple comparisons
<sup>b</sup> Wilcoxon signed rank paired test for paired pre- and post-challenge samples for each individual
<sup>c</sup> Effect size reported as the difference between the two sample means divided by the pooled standard deviation of the log<sub>10</sub> transformed data (Cohen's *d*). The larger the effect size, the larger the difference between the mean of the two sample distributions.

**Figure 1.**
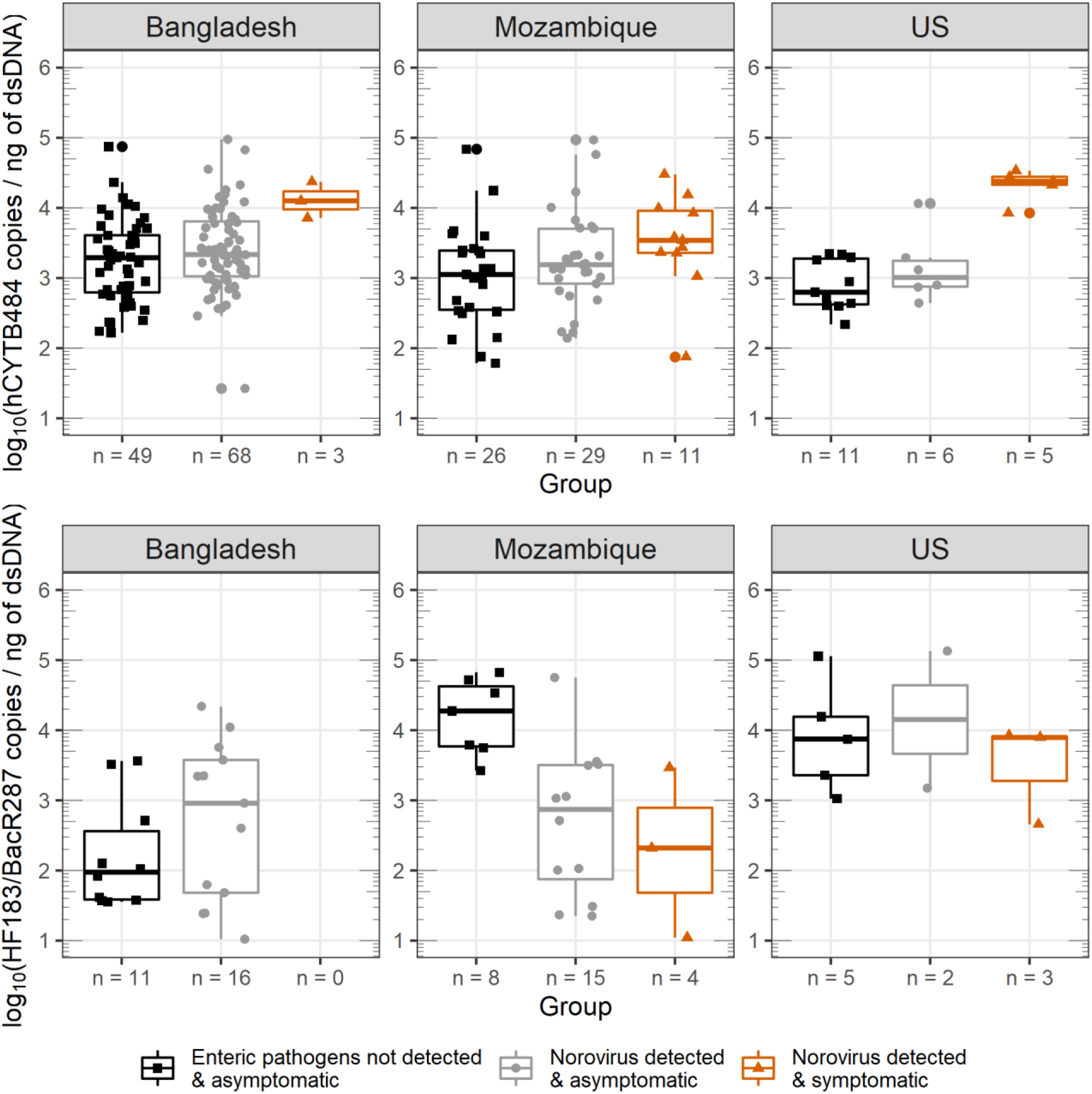
Box and whisker plots of hCYTB484 (top plot) and HF183/BacR287 (bottom plot) concentrations for the various health statuses in this study. Horizontal lines (box) denote the 25^th^, 50^th^, and 75^th^ percentiles and the end of the vertical lines (whiskers) denote the maximum or minimum value of the data that is within 1.5 times the interquartile range over the 75^th^ percentile or under the 25^th^ percentile. All concentrations are plotted as log_10_(concentration + 1) and concentrations from feces are normalized to concentration of dsDNA (ng of dsDNA determined by Qubit). The number of quantifiable samples is shown at x-axis tick mark of each box and whisker (“n = #”). HF183/BacR287 plots have different numbers of quantifiable samples because HF183/BacR287 was detected in only 52% of all samples and quantifiable in 31% of all samples.

The largest increase in concentrations of fecal mtDNA occurred in the US samples, for several possible reasons. Firstly, epithelial cell proliferation declines with age ^23^. Compared to the adults in the US study, the children in the Mozambique and Bangladesh studies may have had higher levels of proliferation even when not experiencing diarrhea. Secondly, fecal mtDNA concentrations may have varied due to environmental enteric dysfunction (EED) ^24^. The Mozambique and Bangladesh study settings had high prevalence of enteric pathogen exposure ^15,18^ as measured by frequency of pathogen detection in feces ^25^: estimated 86% and 88% of feces containing one or more pathogens in Mozambique and Bangladesh samples, respectively. EED, a condition caused by persistent exposure to enteric pathogens, infections, or perturbations and characterized by deleterious changes in the intestinal epithelium, can result in malabsorption and diminished growth and development in children. Despite deleterious changes associated with EED that may potentially change fecal mtDNA concentrations, such as reduced intestinal villi, EED typically presents with few or no acute symptoms, potentially reducing in the effect size we observed in samples from Mozambique and Bangladesh. Lastly, the method of reporting diarrhea differed between the US versus Mozambique and Bangladesh samples: clinical monitoring versus caregiver reported. Caregiver-reported diarrhea is subject to recall biases ^26,27^, which may have resulted in misclassification of samples.

We did not observe any increases in HF183/BacR287 marker copies / ng dsDNA between the enteric^-^asymptomatic and noro^+^symptomatic groups (Figure 1), indicating that the elevated mtDNA concentrations may be specific to mtDNA and not due to a bulk increase in fecal markers. Furthermore, we detected the HF183/BacR287 marker in 52% of samples, with only 31% of samples above the analytical lower limit of quantification (quantifiable), a finding consistent with assessments of HF183 in individual human feces across the globe ^5,28^. In contrast, we quantified 100% of samples in this study for hCYTB484. HF183/BacR287 was quantifiable in less than a third of the samples and did not exhibit any consistent pattern between the enteric^-^asymptomatic and noro^+^symptomatic groups, suggesting that HF183/BacR287 is not widely quantifiable across individual humans nor are levels of HF183/BacR287 concentration indicative of changes in intestinal inflammatory status. For these reasons, these results suggest HF183/BacR287 would be less useful as a biomarker of intestinal inflammation.

A variety of sources and processes related to the health of the gastrointestinal system influence fecal mtDNA concentrations. Because the integrity of the intestinal epithelium is essential to the host’s health, IECs proliferate and are removed in a highly active and regulated cycle ^29^. IEC removal likely depends on the epithelium and host’s health, including various potential mechanisms: engulfment following apoptosis ^30,31^, shedding into the intestinal lumen ^32–35^, shedding in response to pathogen or pathogen-associated insults ^36–38^, and shedding during other pathological states such as inflammatory bowel disease, neoplastic growth ^39^, and wound healing ^34^. Additionally, current evidence of norovirus infection in humans points towards enterocytes in the small intestines as the primary tropism ^40,41^. Noroviruses, as non-enveloped viruses, are presumed to have lytic effects on their host cells ^42^, potentially releasing host mtDNA into the intestinal lumen. Leukocytes can be found in feces from individuals with inflammatory diarrhea ^40,43,44^, and neutrophils are highly abundant first responders, transmigrating across the intestinal epithelium during enteric infections ^45^. Lastly, there is emerging evidence of mtDNA’s immune-signaling role in inflammatory diseases: pathogen-associated signal ^46^ and damage-associated molecular pattern ^13,47^. Many of these processes through which mtDNA is shed in feces are involved with gastrointestinal health, lending plausibility to our observation of increased fecal mtDNA during symptomatic norovirus infections. However, non-pathogenic diseases such as inflammatory bowel disease and neoplastic growth may also cause elevated fecal mtDNA.

Several limitations qualify our results. Prevalences of reported diarrhea in the Bangladesh ^17^ and Mozambique ^15^ trials were low, limiting the number of samples in our analysis and constraining statistical power. Reported diarrhea in these studies was assessed through a caregiver survey and is subject to observational and recall biases ^26,27^. Multiple pathogens were commonly detected in the Bangladesh and Mozambique fecal samples, with norovirus rarely detected alone. Co-infections may have affected fecal mtDNA concentrations as well as symptomology; norovirus may not have always been the cause of symptoms. While norovirus is an important cause of gastroenteritis globally, there are other enteric pathogens that can be transmitted through exposure to fecal contamination in the environment and are relevant to FST.

A human-specific FST marker that is informative of risk of illness is needed because fecal indicator bacteria exhibit non-specificity in the environment (cross-reactivity and regrowth) and because human fecal contamination represents an important risk to human health ^48–52^. In this study, we observed increased concentrations of mtDNA in feces from individuals with symptomatic norovirus infections when compared to feces from individuals without norovirus infections or diarrhea symptoms. This suggests that mtDNA markers may serve as biomarkers of intestinal inflammation and may provide risk-relevant information by increasing in concentration when an individual is at higher risk of transmitting norovirus infection ^53^. However, more work needs to be done to understand current limitations of human mtDNA in FST ^9^. We need better understanding of the cellular sources of fecal mtDNA, intactness of fecal mtDNA after defecation, and what conditions modulate fecal mtDNA concentrations. Approaches investigating processes relevant to gastrointestinal health (e.g., expression of IEC proliferation genes) could also help identify relevant mechanisms that modulate fecal mtDNA. Studies on the fate and persistence of fecal mtDNA ^8,55^ are needed to understand how the signal is attenuated in environments relative to that of infectious pathogens ^56,57^. Before human mtDNA, a nucleic acid marker, can serve as a risk-relevant FST marker, we need to understand its relationship with infectious pathogens ^54^. Because human mtDNA FST markers are typically found at lower concentrations in sewage than those of other human-associated FST markers, improved concentration and recovery methods of mtDNA are needed ^10,58–61^. Better understanding of potential non-fecal sources of mtDNA and at what concentrations non-fecal sources shed mtDNA are also needed ^62^. Potential carry over from consumption of meat or feces of other species should also be investigated ^6,9^. Despite these knowledge and technical gaps, results from this study add to previous evidence supporting the utility of human mtDNA as FST markers.

## Data Availability

All data produced are available in the supplementary information (as an Excel file).

## Acknowledgements

This material is based upon work supported by the National Science Foundation under Grant Number 1511825 and the United States Geological Survey under Project ID 2018GA388B. We received further support from the Bill and Melinda Gates Foundation grant OPP1137224 and the World Bank Strategic Impact Evaluation Fund (SIEF). The funders had no role in study design, data collection and analysis, decision to publish, or preparation of the manuscript. We acknowledge Janet Hatt, Minjae Kim, Angela Peña-Gonzalez, Trent Sumner, Sid Patel, and Victoria Dean.

## For Table of Contents Only

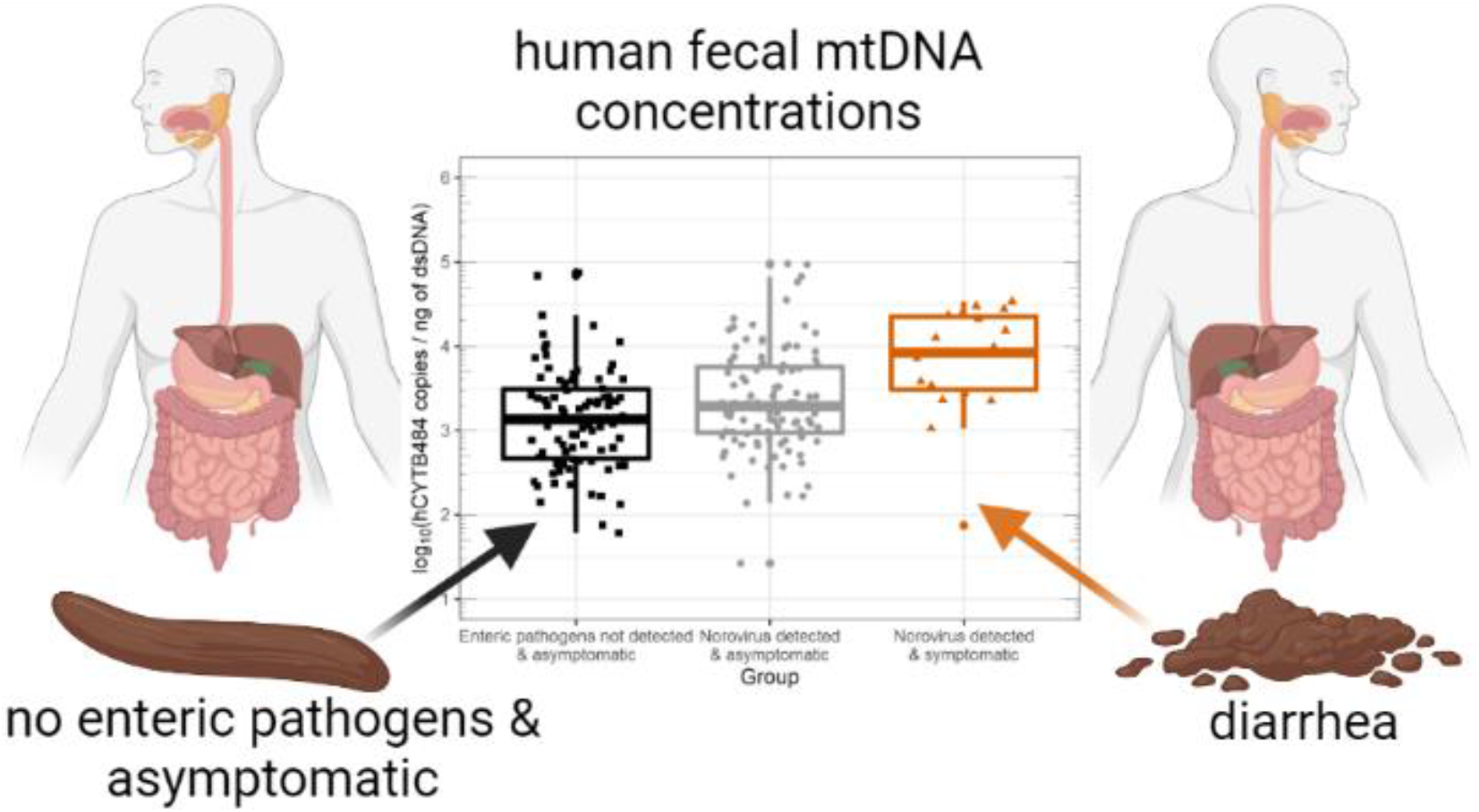

