## Supporting Information for "Elevated fecal mitochondrial DNA from symptomatic norovirus infections suggests potential health relevance of human mitochondrial DNA in fecal source tracking"

**Author Information**

**Kevin J. Zhu** – School of Civil and Environmental Engineering, Georgia Institute of Technology, Atlanta, Georgia 30332, United States

**Brittany Suttner** – School of Civil and Environmental Engineering, Georgia Institute of Technology, Atlanta, Georgia 30332, United States

**Jackie Knee** – Department of Disease Control, London School of Hygiene and Tropical Medicine, London, United Kingdom

**Drew Capone** – Department of Environmental Sciences and Engineering, Gillings School of Global Public Health, University of North Carolina at Chapel Hill, Chapel Hill, North Carolina 27599, United States

**Christine L. Moe** – Center for Global Safe Water, Sanitation, and Hygiene, Rollins School of Public Health, Emory University, Atlanta, Georgia 30322, United States

**Christine E. Stauber** – Department of Population Health Sciences, School of Public Health, Georgia State University, Atlanta, Georgia 30302, United States

**Kostas T. Konstantinidis** – School of Civil and Environmental Engineering, Georgia Institute of Technology, Atlanta, GA 30332, United States

**Thomas E. Wallach** – Division of Pediatric Gastroenterology, SUNY Downstate Health Sciences University, Brooklyn, New York 11203, United States

**Amy J. Pickering** – Department of Civil and Environmental Engineering, University of California, Berkeley, California 94720, United States

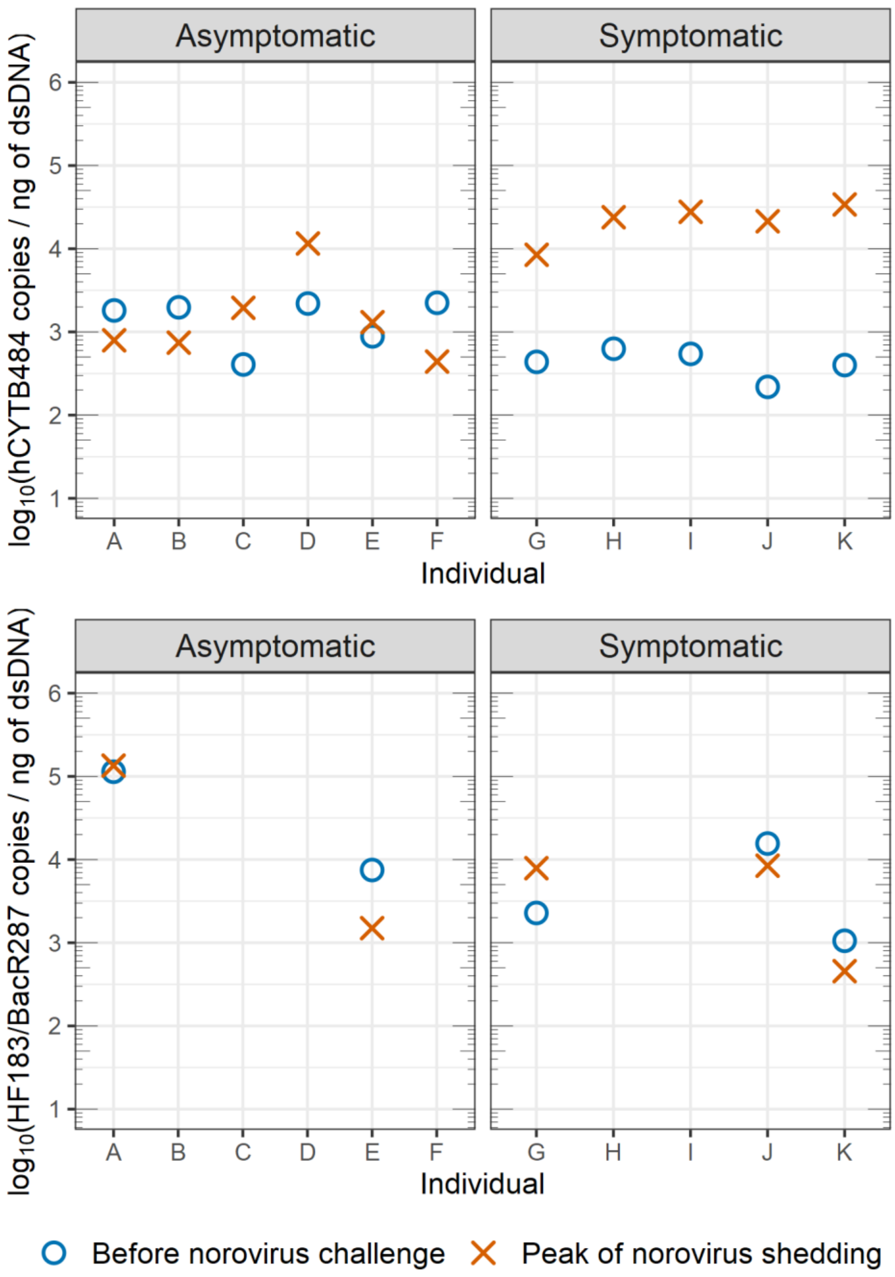


**Figure S1.** hCYTB484 and HF183/BacR287 counts in samples taken from US norovirus challenge study comparing fecal samples taken before the norovirus challenge to fecal samples taken during the peak of norovirus shedding for individuals who became symptomatic or remained asymptomatic for norovirus. Counts of both markers are normalized to ng of dsDNA as determined by Qubit and reported as $\log_{10}$(concentration + 1).

**Table S1.** Comparisons between biological replicates for hCYTB484 and HF183/BacR287 markers showing a mean percent difference of 24% for hCYTB484 and 37% for HF183/BacR287.

| **Sample** | **hCYTB484 Biological Replicate 1 [copies / ng of dsDNA]** | **hCYTB484 Biological Replicate 2 [copies / ng of dsDNA]** | **hCYTB484 Percent Difference [%]** | **HF183/BacR287 Biological Replicate 1 [copies / ng of dsDNA]** | **HF183/BacR287 Biological Replicate 2 [copies / ng of dsDNA]** | **HF183/BacR287 Percent Difference [%]** |
|---|---|---|---|---|---|---|
| 1 | 4.6E+02 | 2.4E+02 | 48 | DBNQ | DBNQ | Not applicable |
| 2 | 2.5E+02 | 2.7E+02 | -9 | DBNQ | DBNQ | Not applicable |
| 3 | 6.8E+02 | 5.7E+02 | 17 | DBNQ | DBNQ | Not applicable |
| 4 | 7.6E+02 | 5.7E+02 | 24 | 4.0E+01 | 2.4E+01 | 40 |
| 5 | 6.8E+02 | 6.1E+02 | 10 | 2.2E+03 | 8.9E+02 | 60 |
| 6 | 5.9E+02 | 8.4E+02 | -43 | DBNQ | DBNQ | Not applicable |
| 7 | 1.8E+03 | 1.2E+03 | 32 | 2.2E+03 | 2.6E+03 | -17 |
| 8 | 2.6E+03 | 1.5E+03 | 42 | No Amplification | No Amplification | Not applicable |
| 9 | 6.0E+03 | 1.9E+03 | 67 | DBNQ | 5.3E-01 | Not applicable |
| 10 | 3.8E+03 | 2.1E+03 | 46 | 1.8E+00 | 5.9E+00 | -230 |
| 11 | 2.0E+03 | 2.1E+03 | -6 | Amplification below aLoD | No Amplification | Not applicable |
| 12 | 3.9E+03 | 2.2E+03 | 42 | Amplification below aLoD | No Amplification | Not applicable |
| 13 | 6.3E+03 | 3.7E+03 | 41 | Amplification below aLoD | Amplification below aLoD | Not applicable |
| 14 | 5.5E+03 | 4.7E+03 | 14 | Amplification below aLoD | No Amplification | Not applicable |
| 15 | 1.4E+04 | 1.0E+04 | 28 | No Amplification | No Amplification | Not applicable |
| Mean | | | 24 (range: -9 to 67) | | | -37 (range: -230 to 60) |

**Table S2.** Minimum Information for Publication of Quantitative Digital PCR Experiments (dPCR MIQE) for the ddPCR materials and methods used in this study.

| ITEM TO CHECK | COMMENT |
|---|---|
| **1. SPECIMEN** | |
| Detailed description of specimen type and numbers | We extracted DNA from archived samples collected in previous studies. Detailed descriptions of specimens can be found elsewhere [1–3]. Number of samples used in this study can be found in Table 1 of the main text. |
| Sampling procedure (including time to storage) | Detailed descriptions of sampling procedures can be found elsewhere [1–3]. |
| Sample aliquotation, storage conditions and duration | All samples were archived at -80°C before DNA extraction for this study. |
| **2. NUCLEIC ACID EXTRACTION** | |
| Description of extraction method including amount of sample processed | We used the MO BIO PowerSoil® kit (Carlsbad, CA, USA) DNA extraction kit for the US samples and the Qiagen QIAamp® 96 PowerFecal QIAcube® HT Kit automated on the Qiagen QIAcube® HT platform (Hilden, Germany) for the Mozambiquan and Bangladeshi samples. |
| Volume of solvent used to elute/resuspend extract | We eluted each sample using 100 µL of elution buffer: Buffer EB for PowerSoil kit and Qiagen Solution C6 for PowerFecal. |
| Number of extraction replicates | We extracted ~7% (n = 15) of the samples in duplicate. See Table S1 for comparisons of extraction replicates. |
| Extraction blanks included? | At least one extraction blank was done with each batch of extractions. No extraction blanks were above the analytical limit of detection. |
| **3. NUCLEIC ACID ASSESSMENT AND STORAGE** | |
| Method to evaluate quality of nucleic acids | We did not assess the quality of nucleic acids. |
| Method to evaluate quantity of nucleic acids (including molecular weight and calculations when using mass) | We used a Qubit 3 Fluorometer (ThermoFisher Scientific, Waltham, MA, USA) with Qubit dsDNA HS Assay Kits to quantify the yield of dsDNA (ng of dsDNA) in each sample. |
| Storage conditions: temperature, concentration, duration, buffer, aliquots | We stored eluted DNA in Buffer ATE (for the PowerFecal extracts) and Buffer C6 (for the PowerSoil extracts) in both diluted and undiluted aliquots for up to 2 years before analyzing on ddPCR at -80°C. |
| Clear description of dilution steps used to prepare working DNA solution | To make any dilutions, we UV-treated microcentrifuge tubes for 20 minutes and made 1 in 10 dilutions using 5 µL of extract and 45 µL of the respective elution buffer (Buffer EB or Solution C6). |

| 4. NUCLEIC ACID MODIFICATION | |
|---|---|
| Template modification (digestion, sonication, pre-amplification, bisulphite etc.) | We did not perform template modification in this study. |
| Details of repurification following modification if performed | We did not perform any repurification following DNA extraction. |
| **5. REVERSE TRANSCRIPTION** | |
| cDNA priming method and concentration | We did not perform reverse transcription in this study. |
| One or two step protocol (include reaction details for two step) | |
| Amount of RNA added per reaction | |
| Detailed reaction components and conditions | |
| Estimated copies measured with and without addition of RT* | |
| Manufacturer of reagents used with catalogue and lot numbers | |
| Storage of cDNA: temperature, concentration, duration, buffer and aliquots | |
| **6. dPCR OLIGONUCLEOTIDES DESIGN AND TARGET INFORMATION** | |
| Sequence accession number or official gene symbol | The two assays we used in this study were: (1) hCYTB484 targeting a human-specific region of the cytochrome *b* gene within the mitchondrial genome and (2) HF183/BacR287 targeting human-associated members of the *Bacteroides* genus. |
| Method (software) used for design and *in silico* verification | We used NCBI BLAST to design and verify *in silico* [4]. |
| Location of amplicon | The hCYTB484 amplicon begins at the 484 bp position of the cytochrome b gene of the revised Cambridge Reference Sequence (reference human mtDNA genome). The HF183/BacR287 amplicon begins at the 180 bp position partial 16S rRNA gene sequence of Bacteroides dorei (accession number AB242143). |
| Amplicon length | The hCYTB484 assay has a length of 121 bp. The HF183/BacR287 assay has a length of 126 bp. |

| | |
|---|---|
| Primer and probe sequences (or amplicon context sequence)** | [5'-3']<br>hCYTB484F: CAATGAATCTGAGGAGGCTAC<br>hCYTB604R: CGTGCAAGAATAGGAGGTG<br>hCYTB520TM: ACCCTCACACGATTCTTTACCTTTCACT<br>HF183: ATCATGAGTTCACATGTCCG<br>BacR287: ATCATGAGTTCACATGTCCG<br>BacR234: ATCATGAGTTCACATGTCCG<br>BacR234IAC: ATCATGAGTTCACATGTCCG |
| Location and identity of any modifications | The hCYTB484 assay had a Zen quencher (Integrated DNA Technologies, Coraville, IA, USA): 5'-/56-FAM/ACC CTC ACA /ZEN/CGA TTC TTT ACC TTT CAC T/3IABkFQ/-3'. The HF183/BacR287 assay had two MGB probes: BacP234MGB: FAM-CTAATGGAACGCATCCC-MGBNFQ and BacP234IAC: HEX-AACACGCCGTTGCTACA-MGBNFQ |
| Manufacturer of oligonucleotides | All primers and probes used in this study were manufactured by IDT (Coraville, IA, USA) except for the MGB probes used in HF183/BacR287. MGB probes were manufactured by Applied Biosystems (Waltham, MA, USA). |
| **7. dPCR PROTOCOL** | |
| Manufacturer of dPCR instrument and instrument model | We used the QX200 droplet digital PCR platform manufactured by Bio-Rad (Hercules, CA, US). |
| Buffer/kit manufacturer with catalogue and lot number | We used BioRad's ddPCR Supermix for Probes (No dUTP) (cat. No. 1863024). We did not record buffer lot numbers for this study. |
| Primer and probe concentration | For hCTYB484: primers at 900 nM and probe at 250 nM. For HF183/BacR287: primers at 1000 nM and probe at 250 nM. |
| Pre-reaction volume and composition (incl. amount of template and if restriction enzyme added) | The pre-reaction total volume (before droplet generation) was 22 µL and composed of 4.99 µL of molecular grade $H_2O$, 11 µL of BioRad ddPCR Supermix for Probes (no dUTP), 0.055 µL of probe, 1.98 µL of primers, and 2 µL of template. We did not add restriction enzymes. |
| Template treatment (initial heating or chemical denaturation) | We did not do any treatments of the template. |
| Polymerase identity and concentration, Mg++ and dNTP concentrations*** | The concentration of divalent cations was 3.8 mM, and the concentration of dNTPs was 0.8 mM. |
| Complete thermocycling parameters | We used 10 min at 95 °C, followed by 40 cycles of 30 s at 95 °C and 60 s at the assay-specific annealing temperature (annealing temperature of 58 °C for HF183/BacR287 and 59 °C for hCYTB484), followed by a 10-min hold at 98 °C. All ramp rates set at 2 °C/s. |

| 8. ASSAY VALIDATION | |
|---|---|
| Details of optimisation performed | For each assay, we conducted a series of experiments to optimize the annealing temperature. First, we ran a temperature gradient spanning approximately 8°C; then, we ran a finer scale temperature gradient (spanning approximately 2°C) by identifying the highest temperatures in which the separation between negative and positive bands reached a limit in the previous gradient. We selected an annealing temperature from the finer scale gradient that gave us the most separation while remaining a relatively high temperature to avoid non-specific amplification. We also experimented with 94°C, 95°C, and 96°C denaturation cycles, finding that 95°C provided the best separation between positive and negative partition signals for the assays used in this study. |
| Analytical specificity (vs. related sequences) and limit of blank (LOB) | Analytical specificity: 97% for hCYTB484 and 80% for HF183/BacR287 [4]. We did not use a limit of blank but instead treated amplification under the analytical limit of detection as "amplification below the analytical limit of detection." We considered amplification below the analytical limit of detection as not detected. |
| Analytical sensitivity/LoD and how this was evaluated | Analytical sensitivity: 100% for hCYTB484 and 51% for HF183/BacR287. This was evaluated by testing 22 cow, 34 pig, 8 chicken, 22 goat, and 222 human feces samples [4]. |
| Testing for inhibitors (from biological matrix/extraction) | We tested for inhibition using the internal amplification control as described in Green et al. 2014 [5]. |
| **9. DATA ANALYSIS** | |
| Description of dPCR experimental design | Detailed description of the dPCR methods can be found in Zhu et al. 2020 [4]. |
| Comprehensive details negative and positive of controls (whether applied for QC or for estimation of error) | We ran at least two wells of UV-treated (for at least 15 minutes) molecular grade water as our negative controls for each plate. For positive controls, we used |
| Partition classification method (thresholding) | Based on previous experience with partitions with intermediate fluorescence, we adopted a moderate approach to partition thresholding. Our method starts with the histogram of the partition fluorescence of each entire plate run. We select the amplitude value for the peak of both the negative and positive bands. We determine the negative and positive bands by comparing with the no template controls and positive controls run on each plate. We then find the midpoint value between the peaks of the positive and negative partition bands and set the threshold at the midpoint value. We argue that this approach has value in fecal source tracking (FST) due to possibilities of detecting degraded target in environmental samples as well as close but not exact sequence matches. |

| | |
|---|---|
| Examples of positive and negative experimental results (including fluorescence plots in supplemental material) | 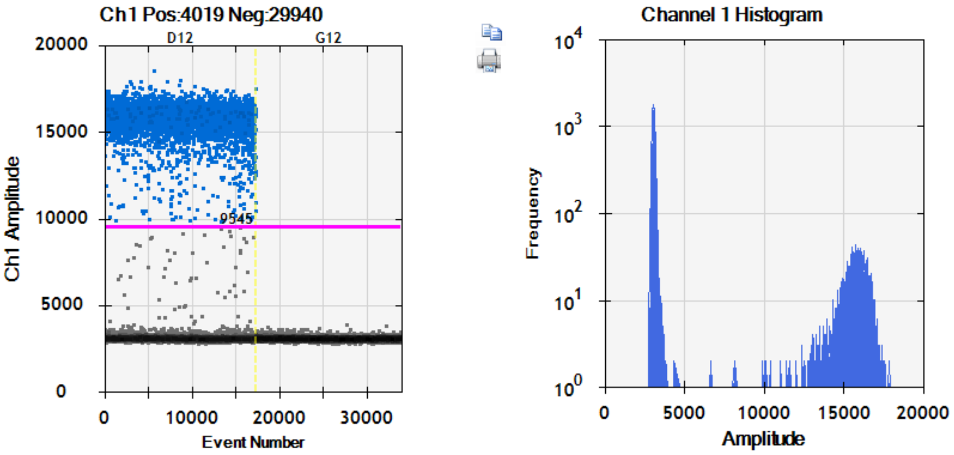  Positive sample (human feces) for hCYTB484 on the left (well labeled D12) showing positive partition band around Channel 1 Amplitude of 15,000 and negative partition band below Channel 1 Amplitude of 5,000. Negative sample (no-template control consisting of UV-treated molecular grade water) on the right (well labeled G12) showing no positive partitions and a negative partition band below Channel 1 Amplitude of 5,000. Note that negative partition bands are similar between the human feces sample and the no-template control. Screenshot taken from Bio-Rad QuantaSoft (Version 1.7.4.01917). |
| Description of technical replication | We performed technical replicates for 25% of the samples. |
| Repeatability (intra-experiment variation) | Coefficient of variation data can be found in Zhu et al. 2020 [4]. |
| Reproducibility (inter-experiment/user/lab etc. variation ) | No evaluation of user or lab variability: only one user analyzed the samples in only one lab. |
| Number of partitions measured (average and standard deviation ) | Mean = 14,113 partitions and standard deviation of 1,665 partitions |
| Partition volume | We did not measure partition volume; instead, we used the assumed volume of 0.85 nL set in Bio-Rad QuantaSoft Version 1.7.4.0917. |
| Copies per partition ($\lambda$ or equivalent ) (average and standard deviation) | Mean $\lambda$ = 0.37. Standard deviation of $\lambda$ = 0.81. |
| dPCR analysis program (source, version) | Bio-Rad QuantaSoft (Version 1.7.4.0917) |
| Description of normalisation method | We normalized to DNA yield by (1) calculating marker copies per µL of extract, (2) quantifying DNA yield per µL of extract via Qubit 3.0 Fluorometer with Qubit High Sensitivity DNA kits (ThermoFisher Scientific, Waltham, MA, USA), and (3) dividing marker copies per µL of extract by ng of dsDNA per µL of extract to obtain marker copies per ng of dsDNA. |
| Statistical methods used for analysis | We applied the Wilcoxon signed rank paired test to the pre- and post-challenge US samples. For the Mozambique and Bangladesh sample sets (cross-sectional data), we used the Kruskal-Wallis test, followed with the Dunn test with Benjamini-Hochberg adjustment. We calculated effect sizes for log10 transformed concentrations through a |

| | |
|---|---|
| | difference in means approach using Cohen's d, the difference between the two means divided by the pooled standard deviation. To compare the relative influences of potential confounders, we fitted a generalized linear model (GLM) using a Gaussian identity function to the Mozambique and Bangladesh sample sets using reported diarrhea and norovirus GI/GII detected/not detected (as determined by the GPP) as the independent variables and log10 values of hCYTB484 normalized to ng of dsDNA as the dependent variable while adjusting for number of pathogens detected (as determined by the GPP), sex, age (continuous, number of months), and study population (Mozambique or Bangladesh). |
| Data transparency | ddPCR results are available in supplementary info as an Excel file. |

**Generalized Linear Model.** We fit a generalized linear model (GLM) to the $\log_{10}$ transformed copies of hCYTB484 normalized to ng of dsDNA using the glm() function in R version 4.0.1. We made the decision to $\log_{10}$ transform hCYTB484 concentrations based off of Box-Cox transformation tests. We fit the GLM to only data from the Mozambiquan and Bangladeshi samples due to the differences in environment (US as a high-income country) and study population (children versus adults) when compared with the US study. Regression coefficient information is shown in Table S3. To assess the fit of the GLM, we plotted the residuals of the GLM using a normal quantile-quantile plot (Figure S2) to assess if the residuals were normally distributed as well as plotted the residuals versus predicted values to assess if the residuals were of constant variance (Figure S3).

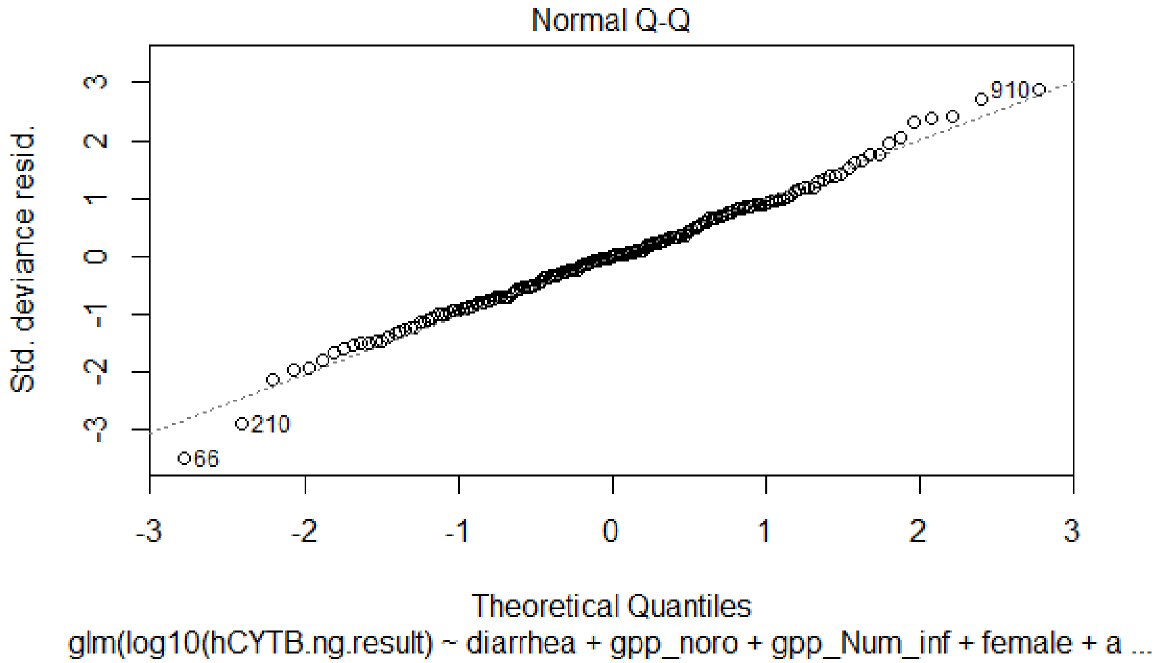


**Figure S2.** A normal quantile-quantile plot of the residuals of the GLM showing linearity, demonstrating that the residuals are normally distributed. Normally distributed residuals of the GLM support the appropriateness of the GLM fit to the data.

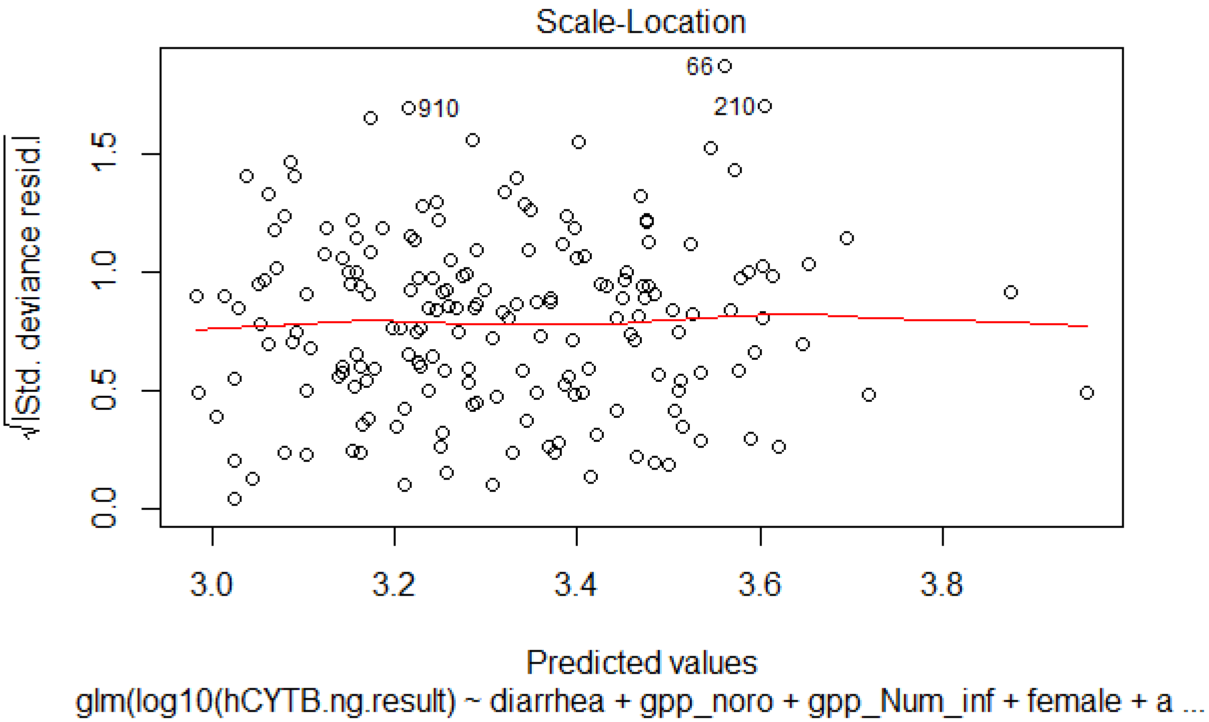


**Figure S3.** A residual by predicted values plot showing that the residuals have approximately constant variance (as indicated by the roughly constant horizontal band pattern as opposed to any curvature pattern), supporting the appropriateness of the GLM fit to the data.

**Table S3.** Coefficients of the generalized linear model (GLM) fitted using only the Bangladesh and Mozambique samples. We fitted the GLM using reported diarrhea and norovirus GI/GII detected/not detected as the independent variables and log10(hCYTB484 copies / ng of dsDNA) as the dependent variable while adjusting for sex, age, and study population.

| | **Intercept** | **Diarrhea** | **Norovirus (Detected / Not detected)** | **Number of GI Infections (GPP)** | **Sex (Male / Female)** | **Age (months)** | **Study Population (Bangladesh / Mozambique)** |
|---|---|---|---|---|---|---|---|
| Estimate | 3.4 | 0.38 | 0.15 | 0.0084 | -0.082 | -0.0052 | -0.26 |
| Std. Error | 0.13 | 0.19 | 0.18 | 0.050 | 0.093 | 0.0033 | 0.11 |
| 95% Confidence Interval Range | 3.14 – 3.66 | 0 – 0.76 | -0.21 – 0.51 | -0.092 – 0.011 | -0.27 – 0.10 | -0.012 – 0.0014 | -0.48 – -0.04 |
| *p*-value | <2e-16 | 0.045 | 0.38 | 0.87 | 0.38 | 0.12 | 0.018 |
| Standardized Regression Coefficient | N/A | 0.16 | 0.12 | 0.023 | -0.064 | -0.12 | 0.19 |
| 95% Confidence Interval Range for Standardized Regression Coefficients | N/A | 0 – 0.32 | -0.16 – 0.39 | -0.25 – 0.03 | -0.21 -- | -0.28 – 0.03 | -0.36 – -0.03 |
